# Optimal symptom combinations to aid COVID-19 case identification: analysis from a community-based, prospective, observational cohort

**DOI:** 10.1101/2020.11.23.20237313

**Authors:** M Antonelli, J Capdevila, A Chaudhari, J Granerod, LS Canas, MS Graham, K Klaser, M Modat, E Molteni, B Murray, CH Sudre, R Davies, A May, LH Nguyen, DA Drew, A Joshi, AT Chan, JP Cramer, T Spector, J Wolf, S Ourselin, CJ Steves, AE Loeliger

**Affiliations:** School of Biomedical Engineering & Imaging Sciences, King’s College London, London, UK; Zoe Global, London, UK; Coalition for Epidemic Preparedness Innovations, London, UK; MRC Unit for Lifelong Health and Ageing at UCL/Centre for Medical Image Computing, Department of Computer Science, UCL, London, UK; Clinical and Translational Epidemiology Unit, Massachusetts General Hospital and Harvard Medical School, Boston, MA, USA; Division of Gastroenterology, Massachusetts General Hospital and Harvard Medical School, Boston, MA, USA; Department of Twin Research and Genetic Epidemiology, King’s College London, London, UK

## Abstract

**Objectives:** Diagnostic work-up following any COVID-19 associated symptom will lead to extensive testing, potentially overwhelming laboratory capacity whilst primarily yielding negative results. We aimed to identify optimal symptom combinations to capture most cases using fewer tests with implications for COVID-19 vaccine developers across different resource settings and public health.

**Methods:** UK and US users of the COVID-19 Symptom Study app who reported new-onset symptoms and an RT-PCR test within seven days of symptom onset were included. Sensitivity, specificity, and number of RT-PCR tests needed to identify one case (test per case [TPC]) were calculated for different symptom combinations. A multi-objective evolutionary algorithm was applied to generate combinations with optimal trade-offs between sensitivity and specificity.

**Findings:** UK and US cohorts included 122,305 (1,202 positives) and 3,162 (79 positive) individuals. Within three days of symptom onset, the COVID-19 specific symptom combination (cough, dyspnoea, fever, anosmia/ageusia) identified 69% of cases requiring 47 TPC. The combination with highest sensitivity (fatigue, anosmia/ageusia, cough, diarrhoea, headache, sore throat) identified 96% cases requiring 96 TPC.

**Interpretation:** We confirmed the significance of COVID-19 specific symptoms for triggering RT-PCR and identified additional symptom combinations with optimal trade-offs between sensitivity and specificity that maximize case capture given different resource settings.

**Highlights:**

- Widely recommended symptoms identified only ∼70% COVID-19 cases
- Additional symptoms increased case finding to > 90% but tests needed doubled
- Optimal symptom combinations maximise case capture considering available resources
- Implications for COVID-19 vaccine efficacy trials and wider public health

## Introduction

Safe and effective vaccines represent the most promising intervention to prevent morbidity and mortality during the coronavirus disease (COVID)-19 pandemic.^1,2^ Positive results have recently emerged from three ongoing vaccine efficacy trials of COVID-19 vaccines.^3-5^ However, further vaccines are required to meet global demand, and vaccines currently in early development may result in better tolerability profiles, scalability, impact on viral shedding, and may be suitable to specific population subgroups. Thus, further important COVID-19 vaccine efficacy trials are predicted to start soon. In a clinical trial, diagnostic testing of suspected cases (e.g., reverse transcription polymerase chain reaction [RT-PCR] for severe acute respiratory syndrome coronavirus 2 [SARS-CoV-2]) could be triggered by the presence of any COVID-19 associated symptom. A household survey in the United Kingdom (UK) showed that fever, cough, anosmia, and ageusia were present on the day of testing in only 60% of symptomatic, RT-PCR positive individuals, implying that other less specific signs/symptoms associated with COVID-19 occur in a substantial number of patients.^6^ The signs/symptoms associated with COVID-19 are extensive and overlap with those of other common viral infections.^7,8^ Thus, diagnostic work-up following any COVID-19 associated symptom may lead to indiscriminate testing and potentially overwhelm laboratory capacity whilst primarily yielding negative results.

Identification of an efficient symptom combination to trigger diagnostic work-up that will capture the majority of COVID-19 cases using the lowest possible number of tests would enable optimum use of laboratory and financial resources in future vaccine efficacy trials. This would also be of wider benefit in public health settings for the early detection of symptomatic SARS-CoV-2 infection. Such data are scant and the triggering symptoms vary between publicly available vaccine efficacy trial protocols.^9-15^

We simulate COVID-19 case finding in a trial population using a community-based, prospective, observational cohort study. Data from UK COVID Symptom Study app ^16^ users were used to quantify how much individual COVID-19 symptoms contribute to COVID-19 case finding and to generate symptom combinations with optimal trade-offs between sensitivity and specificity that maximise the capture of RT-PCR positive cases given different laboratory capacities. The findings were replicated in a dataset of COVID Symptom Study app users in the United States (US).

## Material

### Study design and data source

A community-based cohort study was carried out using data from the COVID Symptom Study app, a free smartphone app launched at the end of March 2020 and developed by Zoe Global (London, UK) in collaboration with King’s College London (London, UK) and Massachusetts General Hospital (Boston, MA, USA).^16^ Users from UK and US report baseline demographic information, data on comorbidities and COVID-19 testing results, and are encouraged to self-report a set of pre-specified symptoms on a daily basis to enable collection of longitudinal information on incident symptoms. This study was approved by the Partners Human Research Committee (Protocol 2020P000909) and King’s College London ethics committee (REMAS ID 18210, LRS-19/20-18210).

### Study population

Individuals were included in the study if they met the following criteria: 1) aged≥18 years, 2) reported developing any symptom between March 24th and September 15th, 2020, and 3) entered a valid RT-PCR test result within the first seven days of symptom onset. App users who recorded a history of COVID-19 were excluded. Data were frozen and extracted on October 21^st^, 2020. UK participants served as a discovery cohort, which was randomly split into training and validation datasets of equal size. US participants served as a replication cohort to confirm the generalisability of the results. Both cohorts were stratified by age (18-54 and ≥55 years) to align with age strata in ongoing COVID-19 vaccine efficacy trials.

## Methods

### Data analyses

Symptoms recorded within three and seven days of symptom onset were included in the analyses (see **Supplementary Table 1** for complete list of symptoms and corresponding questions participants were asked). Analysis of symptoms within the first three days is key to enable testing for SARS-CoV-2 soon after symptom onset while viral load is highest. An additional buffer for inclusion of symptoms within seven days was also used, which may be important to detect development of lower respiratory tract signs indicative of pneumonia. Anosmia and ageusia were considered one symptom in the reporting app.

Participants were classified as symptom-screening positive when they recorded at least one of the symptoms in the symptom combination concerned. This was compared with self-reported RT-PCR results considered the gold standard for COVID-19 case detection. If multiple positive RT-PCR test results were recorded for an individual, only the first was included.

A COVID-19 case was defined as a newly symptomatic individual with a first ever positive RT-PCR test result. For individual symptoms or symptom combinations, three evaluation parameters were considered, taking disease status to be a positive RT-PCR test: 1) sensitivity, computed as the percentage of COVID-19 positive individuals correctly identified, 2) specificity, calculated as the percentage of individuals correctly classified as COVID-19 negative, and 3) the reciprocal of precision, that is the number of RT-PCR tests needed to identify one RT-PCR positive COVID-19 case (i.e. Tests Per Case [TPC]).

### Multi-objective evolutionary optimization

As sensitivity and specificity of a given symptom combination represent conflicting objectives, a multi-objective evolutionary algorithm (MOEA) was used to generate optimal symptom combinations from the data, each characterised by a good trade-off between specificity and sensitivity. Optimisation problems with multiple objectives have a set of optimal solutions (i.e., Pareto-optimal solutions) rather than one single optimal solution. No Pareto-optimal solution is better than the other without further information on the specific objective to be addressed. For MOEA, we employed the well-known NSGAII ^17^ developed in the python package pymoo v0.4.2.1. The optimal set of parameters were derived through experimenting with different values (see **Supplementary Table 2** for parameter information). The training and validation datasets were used to generate and evaluate the Pareto-optimal symptom combinations (referred to as data-inferred symptom combinations).

### Evaluation of individual symptoms and symptom combinations

Sensitivity, specificity, and TPC were evaluated for each individual symptom and symptom combinations using the validation dataset. We considered symptom combinations derived from both clinical experience/guidance (i.e., clinically inferred symptom combinations) and generated from the data using the MOEA (i.e., data-inferred symptom combinations). All evaluations were repeated on the US-replication cohort and on the data stratified by age.

For clinically-inferred symptom combinations we evaluated: 1) respiratory symptoms (cough, dyspnoea), 2) WHO-defined pneumonia symptoms (cough, dyspnoea, fever), 3) COVID-19 specific symptoms as defined by Public Health England (PHE) (fever, cough, dyspnoea, anosmia/ageusia), and 4) extended symptoms (fever, cough, dyspnoea, anosmia/ageusia, fatigue, headache). This latter category was added post-hoc after exploration of the app data indicated high sensitivity of headache and fatigue in other contexts.^18^

Regarding data-inferred symptom combinations, among all the generated combinations, we evaluated the one with highest sensitivity, the one with a sensitivity of ∼90%, and the one characterised by a specificity of ∼50%, which is of interest from a clinical standpoint.

## Results

A total of 122,305 individuals were included in the UK-discovery cohort, of which 1,202 tested COVID-19 positive. In the US-replication cohort, 3,162 individuals were included, of which 79 tested COVID-19 positive. The patient selection flow charts are displayed in **Supplementary Figure 1** and **2. Table 1** shows the demographic characteristics of the population.

**Table 1.** Demographics of study population.

|  | UK-discovery cohort |  | US-replication cohort |  |
| --- | --- | --- | --- | --- |
|  | C-19 RT-PCR positive | C-19 RT-PCR negative | C-19 RT-PCR positive | C-19 RT-PCR negative |
| Total number | 1,202 | 121,103 | 79 | 3,083 |
| Male (%) | 25.1% | 25.3% | 16.0% | 17.5% |
| Mean age, years (SD) | 44.3 (12.5) | 48.5 (13.0) | 52.7 (13.3) | 53.8 (14.7) |
| Mean BMI (SD) | 26.9 (5.75) | 27.3 (5.5) | 27.6 (6.4) | 27.9 (6.0) |

BMI = Body mass index; C-19 = COVID-19; RT-PCR = Reverse transcription polymerase chain reaction; SD = Standard deviation

### Evaluation of individual symptoms

The sensitivity, specificity, and TPC for each individual symptom reported within three and seven days of symptom onset are displayed in **Table 2**. Using the UK-discovery cohort, the individual symptoms with the highest sensitivity in both three- and seven-day analyses were headache and fatigue (67% and 65% for three-day analysis and 75% and 78% for seven-day analyses). Similar results were obtained with data from the US-replication cohort and when data were stratified by age. The sensitivity of anosmia/ageusia in the UK-discovery cohort was only 22% and 49% in the three- and seven days analyses, respectively. Anosmia/ageusia, however, had the lowest TPC (20 and 10 for three- and seven-day analyses, respectively).

**Table 2.** Sensitivity, specificity, and TPC for each individual symptom computed on the UK-discovery cohort.

| Symptom | Age group | 3-day analysis |  |  |  |  |  | 7-day analysis |  |  |  |  |  |
| --- | --- | --- | --- | --- | --- | --- | --- | --- | --- | --- | --- | --- | --- |
|  |  | Sensitivity (%) |  | Specificity (%) |  | TPC |  | Sensitivity (%) |  | Specificity (%) |  | TPC |  |
|  |  | UK | US | UK | US | UK | US | UK | US | UK | US | UK | US |
| Headache | All | 66.8 | 70.9 | 52.4 | 49.7 | 76 | 30 | 75.6 | 81.0 | 48.3 | 45.4 | 70 | 29 |
|  | [18-54] | 67.8 | 73.8 | 50.7 | 48.6 | 67 | 27 | 76.9 | 83.3 | 46.2 | 43.1 | 62 | 27 |
|  | [55+] | 63.1 | 67.6 | 55.8 | 50.7 | 111 | 34 | 71.2 | 78.4 | 52.6 | 47.5 | 102 | 31 |
| Fatigue | All | 64.9 | 73.4 | 53.7 | 47.2 | 76 | 31 | 77.8 | 87.3 | 49.7 | 42.8 | 66 | 28 |
|  | [18-54] | 64.2 | 71.4 | 53.5 | 49.5 | 66 | 28 | 76.9 | 92.9 | 49.2 | 44.6 | 58 | 24 |
|  | [55+] | 67.5 | 75.7 | 53.9 | 45.1 | 108 | 34 | 81.2 | 81.1 | 50.7 | 41.0 | 93 | 34 |
| Sore throat | All | 47.3 | 36.7 | 59.1 | 59.1 | 92 | 47 | 54.8 | 49.4 | 55.8 | 55.9 | 82 | 38 |
|  | [18-54] | 48.6 | 45.2 | 56.1 | 53.8 | 83 | 40 | 55.3 | 54.8 | 52.4 | 49.9 | 76 | 36 |
|  | [55+] | 42.9 | 27.0 | 65.4 | 64.2 | 127 | 60 | 53.1 | 43.2 | 62.9 | 61.6 | 107 | 41 |
| Persistent cough | All | 35.9 | 55.7 | 86.3 | 76.5 | 41 | 18 | 43.4 | 65.8 | 84.6 | 73.1 | 37 | 18 |
|  | [18-54] | 35.8 | 50.0 | 85.6 | 78.2 | 37 | 18 | 42.9 | 61.9 | 83.8 | 74.4 | 34 | 17 |
|  | [55+] | 36.1 | 62.2 | 87.6 | 74.9 | 55 | 19 | 45.4 | 70.3 | 86.2 | 71.8 | 47 | 19 |
| Fever | All | 35.3 | 34.2 | 88.9 | 86.6 | 34 | 17 | 44.8 | 49.4 | 87.0 | 83.4 | 30 | 15 |
|  | [18-54] | 35.8 | 35.7 | 88.4 | 86.5 | 30 | 15 | 45.0 | 47.6 | 86.3 | 82.6 | 27 | 15 |
|  | [55+] | 33.3 | 32.4 | 89.9 | 86.7 | 48 | 19 | 44.2 | 51.4 | 88.4 | 84.1 | 41 | 15 |
| Myalgia | All | 32.2 | 43.0 | 86.1 | 82.9 | 46 | 17 | 43.8 | 59.5 | 84.2 | 79.6 | 37 | 15 |
|  | [18-54] | 32.8 | 42.9 | 86.2 | 85.1 | 39 | 14 | 44.8 | 61.9 | 84.2 | 81.8 | 32 | 12 |
|  | [55+] | 30.2 | 43.2 | 85.7 | 80.8 | 75 | 21 | 40.4 | 56.8 | 84.1 | 77.6 | 61 | 19 |
| Hoarse voice | All | 23.7 | 31.6 | 89.9 | 88.0 | 46 | 17 | 33.7 | 44.3 | 88.0 | 84.8 | 37 | 15 |
|  | [18-54] | 23.1 | 33.3 | 89.9 | 87.6 | 41 | 15 | 33.4 | 40.5 | 87.8 | 84.4 | 33 | 16 |
|  | [55+] | 25.8 | 29.7 | 90.0 | 88.4 | 62 | 19 | 34.6 | 48.6 | 88.4 | 85.2 | 52 | 15 |
| Skipped meals | All | 22.8 | 34.2 | 88.9 | 80.1 | 52 | 25 | 33.0 | 57.0 | 87.5 | 77.0 | 39 | 18 |
|  | [18-54] | 22.5 | 38.1 | 89.1 | 82.5 | 45 | 18 | 32.4 | 54.8 | 87.5 | 79.3 | 35 | 15 |
|  | [55+] | 23.8 | 29.7 | 88.6 | 77.8 | 76 | 34 | 35.4 | 59.5 | 87.5 | 74.8 | 55 | 20 |
| Chest pain | All | 22.5 | 21.5 | 89.1 | 86.3 | 52 | 27 | 33.7 | 32.9 | 87.4 | 83.0 | 39 | 22 |
|  | [18-54] | 23.1 | 14.3 | 88.6 | 85.9 | 46 | 38 | 34.3 | 28.6 | 86.6 | 81.3 | 35 | 26 |
|  | [55+] | 20.6 | 29.7 | 90.1 | 86.7 | 76 | 21 | 31.5 | 37.8 | 88.9 | 84.5 | 54 | 19 |
| Anosmia/ageusia | All | 21.8 | 13.9 | 96.1 | 95.7 | 20 | 14 | 48.7 | 46.8 | 95.4 | 94.7 | 10 | 6 |
|  | [18-54] | 22.9 | 9.5 | 95.8 | 95.4 | 17 | 19 | 51.3 | 47.6 | 95.0 | 94.2 | 9 | 6 |
|  | [55+] | 17.5 | 18.9 | 96.7 | 96.0 | 30 | 10 | 39.2 | 45.9 | 96.2 | 95.1 | 16 | 6 |
| Dyspnoea | All | 20.4 | 22.8 | 89.9 | 86.1 | 53 | 26 | 32.3 | 39.2 | 88.0 | 83.1 | 38 | 19 |
|  | [18-54] | 21.3 | 19.0 | 90.0 | 86.3 | 43 | 28 | 33.3 | 40.5 | 87.9 | 82.5 | 32 | 17 |
|  | [55+] | 17.1 | 27.0 | 89.8 | 85.9 | 95 | 24 | 28.5 | 37.8 | 88.3 | 83.7 | 64 | 20 |
| Diarrhoea | All | 19.1 | 19.0 | 82.5 | 76.8 | 97 | 51 | 27.1 | 38.0 | 80.3 | 72.9 | 74 | 30 |
|  | [18-54] | 19.7 | 16.7 | 82.5 | 76.9 | 81 | 53 | 28.0 | 38.1 | 80.1 | 72.7 | 63 | 28 |
|  | [55+] | 16.7 | 21.6 | 82.5 | 76.6 | 165 | 50 | 23.8 | 37.8 | 80.8 | 73.0 | 123 | 33 |
| Abdominal pain | All | 14.1 | 16.5 | 83.4 | 82.2 | 124 | 45 | 21.3 | 31.6 | 81.3 | 79.5 | 90 | 28 |
|  | [18-54] | 13.6 | 19.0 | 83.6 | 84.1 | 110 | 33 | 21.7 | 31.0 | 81.3 | 81.3 | 76 | 24 |
|  | [55+] | 15.9 | 13.5 | 82.9 | 80.4 | 169 | 66 | 20.0 | 32.4 | 81.2 | 77.9 | 143 | 32 |
| Delirium | All | 8.5 | 12.7 | 92.4 | 89.9 | 95 | 34 | 13.5 | 26.6 | 91.2 | 87.8 | 66 | 20 |
|  | [18-54] | 8.3 | 14.3 | 92.9 | 90.4 | 79 | 26 | 13.4 | 21.4 | 91.7 | 87.7 | 55 | 23 |
|  | [55+] | 9.1 | 10.8 | 91.4 | 89.4 | 148 | 45 | 13.8 | 32.4 | 90.4 | 87.8 | 107 | 18 |

TPC = Tests per case

These results are confirmed by **Figure 1**, which displays the frequency of the symptoms for the UK-discovery cohort for both COVID-19 positive and negative cases.

**Figure 1.**
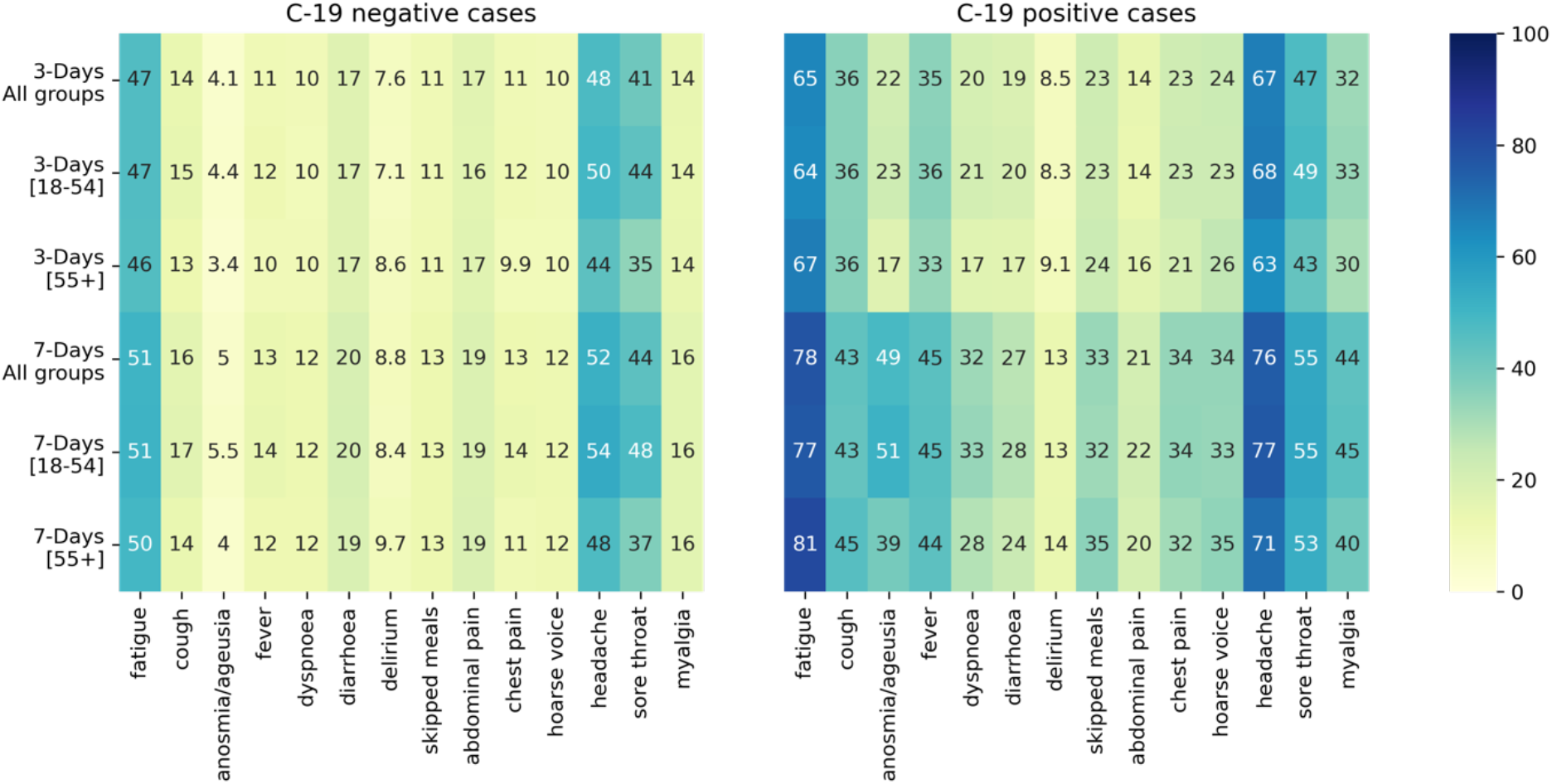
Symptom frequency for COVID-19 negative (left) and COVID-19 positive (right) cases.

### Evaluation of symptom combinations

The sensitivity, specificity, and TPC of both clinically- and data-inferred symptom combinations, computed on the UK-validation and US-replication cohorts, and reported within three and seven days of symptom onset are displayed in **Table 3**.

**Table 3.** Sensitivity, specificity, and TPC for the clinically and data-inferred combinations of symptoms, computed on the held-out validation dataset.

|  |  |  | Three-day analysis |  |  |  |  |  | Seven-day analysis |  |  |  |  |  |
| --- | --- | --- | --- | --- | --- | --- | --- | --- | --- | --- | --- | --- | --- | --- |
|  |  |  | Sensitivity (%) |  | Specificity (%) |  | TPC |  | Sensitivity (%) |  | Specificity (%) |  | TPC |  |
|  | Symptom | Age group | UK | US | UK | US | UK | US | UK | US | UK | US | UK | US |
| Clinically inferred symptoms | Respiratory symptoms<sup>1</sup> | All | 46.4 | 48.1 | 81.9 | 76.6 | 42 | 21 | 58.1 | 64.6 | 79.1 | 72.3 | 37 | 19 |
|  |  | [18-54] | 47.1 | 45.2 | 81.6 | 76.8 | 37 | 20 | 58.5 | 64.3 | 78.5 | 71.6 | 33 | 18 |
|  |  | [55+] | 44.3 | 51.4 | 82.5 | 76.5 | 60 | 22 | 56.7 | 64.9 | 80.4 | 73.0 | 51 | 20 |
|  | WHO defined pneumonia<sup>2</sup> | All | 59.8 | 74.7 | 71.7 | 59.5 | 51 | 23 | 71.4 | 84.8 | 68.4 | 54.5 | 46 | 23 |
|  |  | [18-54] | 59.9 | 69.0 | 71.0 | 61.3 | 46 | 22 | 70.7 | 83.3 | 67.4 | 55.5 | 42 | 21 |
|  |  | [55+] | 59.5 | 81.1 | 73.2 | 57.8 | 68 | 24 | 73.9 | 86.5 | 70.6 | 53.6 | 59 | 25 |
|  | C-19-specific symptoms<sup>3</sup> | All | 69.0 | 79.7 | 69.6 | 57.6 | 47 | 23 | 83.7 | 92.4 | 66.2 | 52.6 | 42 | 22 |
|  |  | [18-54] | 69.5 | 73.8 | 68.8 | 59.5 | 42 | 22 | 84.4 | 92.9 | 65.1 | 53.8 | 37 | 20 |
|  |  | [55+] | 67.2 | 86.5 | 71.4 | 55.7 | 64 | 24 | 81.3 | 91.9 | 68.7 | 51.4 | 57 | 25 |
|  | Extended symptoms<sup>4</sup> | All | 92.0 | 96.2 | 25.9 | 21.1 | 85 | 35 | 96.7 | 98.7 | 22.9 | 17.9 | 81 | 35 |
|  |  | [18-54] | 92.6 | 95.2 | 25.0 | 22.2 | 76 | 32 | 96.6 | 100.0 | 21.7 | 18.6 | 72 | 32 |
|  |  | [55+] | 90.1 | 97.3 | 27.9 | 20.1 | 120 | 38 | 97.0 | 97.3 | 25.6 | 17.3 | 112 | 39 |
| Data-inferred subsets | Combination with highest sensitivity<sup>5</sup> | All | 96.3 | 96.2 | 11.9 | 9.8 | 96 | 40 | 99.2 | 98.7 | 10.4 | 8.2 | 92 | 39 |
|  |  | [18-54] | 96.9 | 95.2 | 10.4 | 8.0 | 85 | 38 | 99.4 | 100 | 8.8 | 6.6 | 80 | 36 |
|  |  | [55+] | 94.4 | 97.2 | 15.2 | 11.6 | 141 | 42 | 98.5 | 97.3 | 13.7 | 9.8 | 134 | 90 |
|  | Combination with sensitivity ~ 90%<sup>6</sup> | All | 92.2 | 96.2 | 22.4 | 15.6 | 89 | 36 | 94.7 | 96.2 | 37.8 | 29.3 | 68 | 31 |
|  |  | [18-54] | 92.7 | 95.2 | 21.5 | 19.2 | 78 | 33 | 93.2 | 100 | 37.1 | 31.3 | 59 | 27 |
|  |  | [55+] | 91.3 | 97.3 | 23.9 | 17.9 | 131 | 39 | 97.7 | 97.3 | 26.8 | 18.8 | 115 | 38 |
|  | Combination with specificity ~ 50%<sup>7</sup> | All | 76.4 | 84.8 | 40.9 | 40.0 | 72 | 30 | 87.3 | 91.1 | 49.2 | 38.9 | 59 | 29 |
|  |  | [18-54] | 76.5 | 80.9 | 48.0 | 42.5 | 63 | 28 | 88.7 | 92.9 | 49.6 | 39.9 | 50 | 25 |
|  |  | [55+] | 79.3 | 89.2 | 49.0 | 37.8 | 101 | 32 | 82.3 | 89.2 | 50.7 | 37.9 | 92 | 32 |

<sup>1</sup>Cough, dyspnoea; <sup>2</sup>Cough, dyspnoea, fever; <sup>3</sup>Fever, cough, dyspnoea, and anosmia/ageusia; <sup>4</sup>Fever, cough, dyspnoea, anosmia/ageusia, fatigue, and headache; <sup>5</sup>Fatigue, anosmia/ageusia, persistent cough, diarrhoea, headache and sore throat; <sup>6</sup>(3-day) Fatigue, anosmia/ageusia, persistent cough, dyspnoea, diarrhoea, headache, (7-day) fatigue, fever, anosmia/ageusia, persistent cough; <sup>7</sup> (3-day) Fatigue, fever, anosmia/ageusia, (7-day) Anosmia/ageusia, persistent cough, dyspnoea, diarrhoea, skipped meals, myalgia TPC = Tests per case

Cough or dyspnoea were reported by 46% of individuals positive for COVID-19 within the first three days of symptom onset. The addition of fever (i.e., WHO-defined pneumonia symptom combination) increased sensitivity to 60%, while the further addition of anosmia/ageusia (i.e., PHE COVID-19 specific symptom combination) increased sensitivity to 69%. When headache and fatigue are added, (i.e., extended symptom combination) the proportion of COVID-19 cases identified increased to 92% but the TPC doubled compared to the respiratory symptom combination (42 versus 85). Similarly, within seven days of symptom onset, COVID-19 specific and extended symptom combination were reported in 84% and 97% of RT-PCR positive cases, at the cost of 42 and 81 TPC, respectively. Similar results were obtained when data were stratified by age. The sensitivity estimates from the US-replication cohort were higher for all four combinations; extended symptom combination estimates reached 96% and 99% for the three- and seven-day analyses, respectively. On the contrary, the specificity decreased to 21% and 18%, although TPC values were lower for the US-replication cohort.

Among data-inferred symptom combinations, the one with highest sensitivity (fatigue, anosmia/ageusia, cough, diarrhoea, headache, and sore throat) identified 96% and 99% of RT-PCR positive COVID-19 cases and required 96 and 92 TPC in the three- and seven-day analyses, respectively. The sensitivity results were similar for the US-replication cohort and by age. However, the number of tests needed for those aged ≥55 years increased by 30% for both the three-day and seven-day analyses.

**Figure 2** displays the three data-inferred symptom combinations for both three- and seven-day analyses. Anosmia/ageusia were included in all three symptom combinations at both time points, fatigue was included in all symptom combinations for the three-day analyses, and cough for the seven-day analyses. Headache was slightly more important when symptoms were recorded within three days of onset. Diarrhoea as an individual symptom was not predictive of a positive COVID-19 RT-PCR result but became predictive when associated with other symptoms.

All the Pareto-optimal symptom combinations generated by the MOEA are displayed in **Figure 3**. Each point (solution) of the Pareto corresponds to a certain symptom combination with a related sensitivity, specificity, and TPC (see **Supplementary Table 4** and **5** for the complete list of solutions for three- and seven-day analyses, respectively). These generated symptom combinations achieved similar values of sensitivity and specificity for the UK-training, UK-validation, and US-replication cohorts, thus confirming the validity of this methodology. Moreover, results were also confirmed for the two age groups.

**Figure 2.**
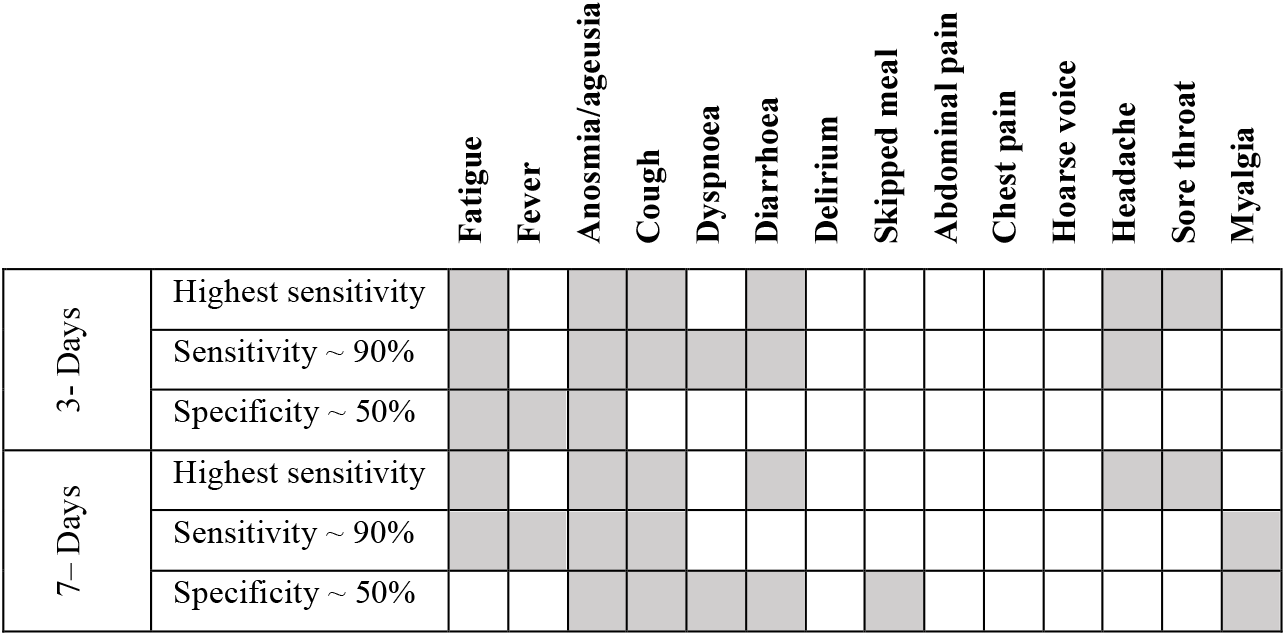
Combination of symptoms with highest sensitivity, sensitivity ∼ 90%, and specificity ∼50%.

**Figure 3.**
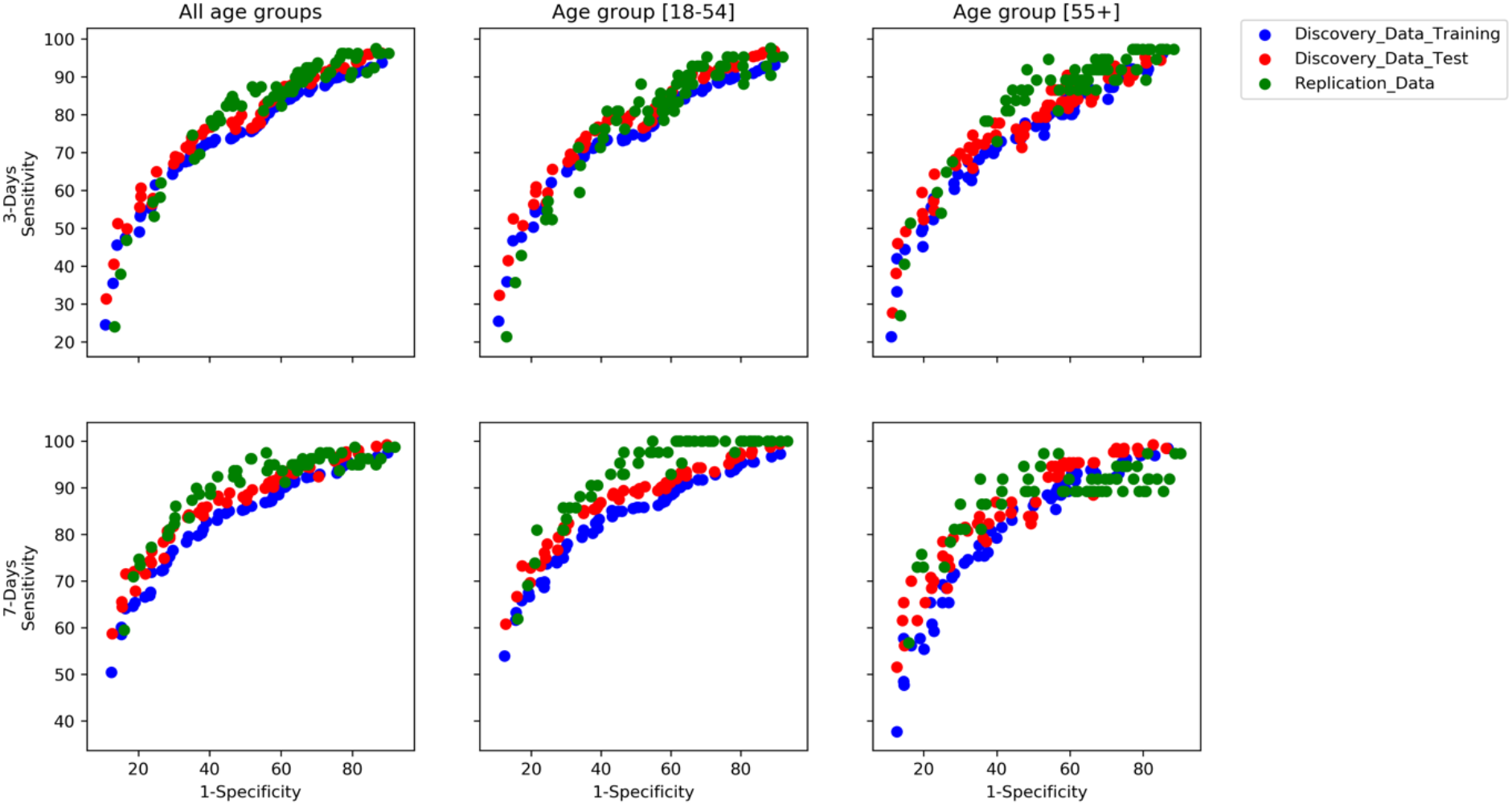
Pareto of optimal subset generated by the multi-objective evolutionary algorithm for three- and seven-day analyses.

**Figure 4** displays the frequency of symptoms selected in symptom combinations with a sensitivity ≥90%. Fatigue, cough, and anosmia/ageusia were present in most symptom combinations with high specificity. Diarrhoea was selected ∼60% of the time for the three-day analyses.

**Figure 4.**
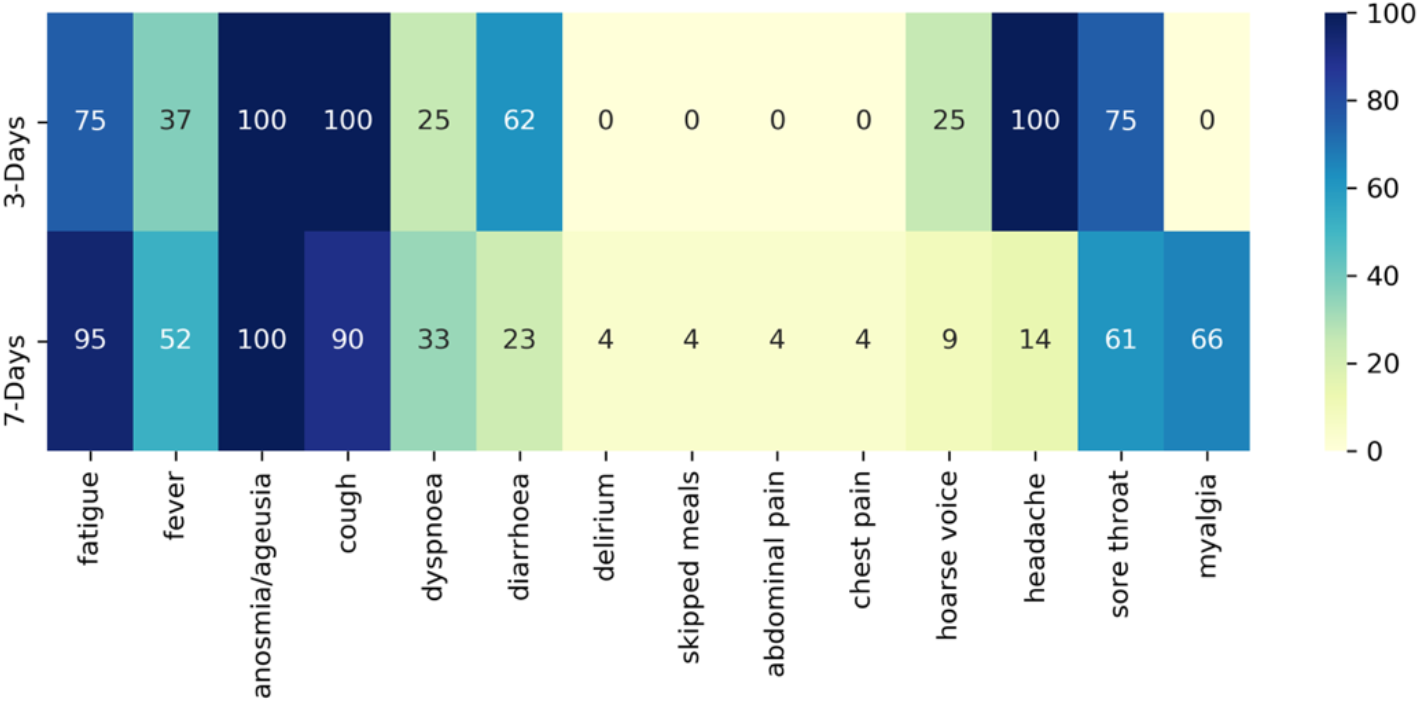
Percentage of a symptom’s appearance in symptom combinations with sensitivity ≥90 %.

Each point represents a subset of symptoms characterised by a different trade-off between sensitivity and specificity.

## Discussion

We present data from, what is to our knowledge, the largest community-based COVID-19 symptom cohort study with the aim to quantify the contribution of various symptoms and symptom combinations associated with COVID-19 to RT-PCR positive case-finding. COVID-19 symptoms and RT-PCR test results were collected prospectively which allowed us to select newly symptomatic individuals and simulate a clinical trial situation in which RT-PCR tests are typically conducted within three days after symptom onset. We confirm the significance of symptoms (fever, cough, anosmia/ageusia) widely considered important for triggering a RT-PCR test and extend this to include additional symptoms (fatigue, sore throat, headache, diarrhoea). The proposed approach enables the selection of symptom combinations to maximise the capture of cases without overwhelming laboratory capacity. Our findings may help to optimise the choice of triggering symptoms for diagnostic work-up in COVID-19 vaccine efficacy trials or in a wider public health setting.

In an efficacy trial, it is important to capture all COVID-19 cases with pulmonary involvement as signs/symptoms of pneumonia define moderate or severe COVID-19. Therefore, the signs/symptoms that characterise WHO-defined COVID-19 pneumonia (fever, cough, dyspnoea, tachypnoea) should trigger diagnostic work-up in a trial participant.^19^ Additionally, anosmia/ageusia have the highest sensitivity of all reported COVID-19 symptoms.^9,20^ Although our findings support the inclusion of these COVID-19 specific symptoms, they also show that this combination correctly identified only 69% and 83% of COVID-19 cases in the three- and seven-days analyses. This has important implications in terms of cases missed as the COVID-specific symptoms align with the current PHE definition of a possible COVID-19 case.^21^ We found that the addition of headache and fatigue (i.e., extended symptoms) increased the proportion of COVID-19 cases correctly identified to 92% but also almost doubled the TPC (from 47 to 85). Thus, an increase in sensitivity comes at a cost.

Application of MOEA identified fatigue, anosmia/ageusia, cough, diarrhoea, headache, and sore throat as the symptom combination with the highest sensitivity in three- and seven-day analyses. Diarrhoea and sore throat were identified as symptoms that may increase case finding in an efficient way, in addition to those symptoms already considered important for triggering an RT-PCR test. In situations where there is a limited testing capacity, we provide a range of optimal symptom combinations that could be used, given different target numbers of TPC identified. This finding may prove useful for COVID-19 vaccine developers or in public health settings when deciding which symptoms should trigger testing to optimise financial and logistical resource utilisation. Importantly, all the symptoms that constitute the combination with the highest sensitivity have been included as triggering symptoms in publicly available clinical trial protocols of ongoing vaccine efficacy trials.^9-14^

Few studies have been published that assess COVID-19 symptoms in community-based cohorts. Menni et al. presented results using data generated from this COVID-19 Symptom Study app; however, the aim was different and only data from March-April 2020 were included.^22^ We extend these data to September 2020 and, importantly, consider the results from the perspective of a potential COVID-19 vaccine developer. Menni et al. suggest anosmia/ageusia, fatigue, persistent cough, and loss of appetite might together identify individuals with COVID-19.^22^ A separate COVID-19 symptom app from Germany suggests nausea and vomiting have a stronger predictive value for COVID-19 infection than symptoms such as sore throat or persistent cough.^23^ Thus, both studies identify gastrointestinal symptoms as important in identifying cases of COVID-19. Our study reports similar findings with diarrhoea found to be important to case finding. More recently, in another community-based observational study, sensitivity, specificity, and positive and negative predictive values were reported for retrospectively collected symptoms and symptom combinations that occurred during the 14-day period prior to screening for SARS-CoV-2 infection in a US seroprevalence study.^24^ The two symptom clusters most associated with SARS-CoV-2 infection were: 1) ageusia, anosmia, and fever, and 2) shortness of breath, cough, and chest pain. In our study, dyspnoea was rarely and chest pain never selected as part of an efficient symptom combination likely due to dyspnoea often occurring later in the disease course.^25^ The sensitivity of dyspnoea increased in the seven-day compared to three-day analyses. However, the importance of dyspnoea as a symptom of pulmonary involvement makes it a critical triggering symptom in vaccine efficacy trials. Tachypnoea, which is included in the WHO-defined definition for pneumonia, was not captured as a symptom in the app per se; however, it likely co-occurs with dyspnoea. Headache and diarrhoea were more likely to be selected in the three-day scenario and fever during the seven-day scenario again, reflecting different timings of symptoms in the disease course.

The sensitivity of symptoms and various clinically inferred symptom combinations were similar for the age groups (18-54 and ≥55 years); however, the TPC was higher in the ≥55 years age group. This suggests self-reporting may work better for younger than older individuals. The sensitivity, specificity, and TPC computed on the US-replication cohort were higher than for the UK-discovery cohort possibly due to different testing practices and public health measures adopted in each country. It will be important for these findings to be validated in low- and middle-income country (LMIC) settings as COVID-19 vaccine efficacy trials are likely to be conducted in high income countries as well as LMICs. Vaccine developers should take into account regional considerations such as background incidence of co-infection and other trial-related aspects when interpreting these results.

This study has many strengths, including the large sample size and cost-effectiveness of the data source. Also, our study is community-based and adds important data as most studies that have assessed symptoms in COVID-19 have involved hospital-based populations. Some limitations, however, also need consideration. First, the results are based on data self-reported through a mobile app and therefore biased towards people with smartphone access. However, the app included a feature to enable reporting on behalf of someone else given their consent. Second, reported test results were not externally verified, however, antigen tests were not available during the study period, thus minimising risk of participant confusion regarding precise swab tests. As the precise RT-PCR used was not recorded and likely varied between participants, false positive rates were not known and results taken at face value. A further limitation is that app users may not be representative of the wider population. Finally, these data were generated in the spring and summer months when the incidence of concurrent respiratory infections (e.g., influenza) is low. The latter may have implications for trials conducted in winter.

In summary, we confirm the significance of symptoms widely recommended for triggering RT-PCR and identified additional symptom combinations to enable efficient trade-off between the number of positive cases detected and tests needed. Our findings may help optimise the choice of triggering symptoms for diagnostic work-up in COVID-19 vaccine efficacy trials and also have wider public health implications.

## Data Availability

Data collected in the COVID-19 Symptom Study smartphone application are being shared with other health researchers through the UK National Health Service-funded Health Data Research UK (HDRUK) and Secure Anonymised Information Linkage consortium housed in the UK Secure Research Platform (Swansea, UK). Anonymised data are available to be shared with HDRUK researchers according to their protocols in the public interest (https://web.www.healthdatagateway.org/dataset/fddcb382-3051-4394-8436-b92295f14259). US investigators are encouraged to coordinate data requests through the Coronavirus Pandemic Epidemiology Consortium (https://www.monganinstitute.org/cope-consortium).

## Acknowledgements

Zoe provided in kind support for all aspects of building, running and supporting the app and service to all users worldwide. CEPI provided funding for the analysis of the data. Support for this study was provided by the NIHR-funded Biomedical Research Centre based at GSTT NHS Foundation Trust. Investigators also received support from the Wellcome Trust, the MRC/BHF, Alzheimer’s Society, EU, NIHR, CDRF, and the NIHR-funded BioResource, Clinical Research Facility and BRC based at GSTT NHS Foundation Trust in partnership with KCL, the UK Research and Innovation London Medical Imaging & Artificial Intelligence Centre for Value Based Healthcare, the Wellcome Flagship Programme (WT213038/Z/18/Z), the Chronic Disease Research Foundation, and DHSC. DAD is supported by the National Institute of Diabetes and Digestive and Kidney Diseases K01DK120742 and by the American Gastroenterological Association AGA-Takeda COVID-19 Rapid Response Research Award (AGA2021-5102). ATC was supported in this work through a Stuart and Suzanne Steele MGH Research Scholar Award. The Massachusetts Consortium on Pathogen Readiness (MassCPR) and Mark and Lisa Schwartz supported MGH investigators (LHN, DAD, ADJ, ATC).

## Supplementary tables

**Supplementary Table 1.** List of self-reported symptoms and corresponding question used in the reporting app.

| Symptoms | Question |
| --- | --- |
| Fatigue | Are you experiencing unusual fatigue? |
| Anosmia/ageusia | Do you have a loss of smell/taste? |
| Cough | Do you have a persistent cough |
| Dyspnoea | Are you experiencing unusual shortness of breath? |
| Diarrhoea | Are you experiencing diarrhoea? |
| Delirium | Do you have any of the following symptoms: confusion, disorientation, or drowsiness? |
| Skipped meals | Have you been skipping meals? |
| Abdominal pain | Do you have an unusual abdominal pain? |
| Chest pain | Are you feeling an unusual chest pain or tightness in your chest? |
| Hoarse voice | Do you have an unusually hoarse voice? |
| Headache | Do you have a headache? |
| Sore throat | Do you have a sore throat? |
| Myalgia | Do you have unusual strong muscle pains? |

**Supplementary Table 2.** NSGAII parameters.

| Parameter | Value |
| --- | --- |
| Population size | 80 |
| Number of offspring | 40 |
| Total number of generations | 500 |
| Parent selection | Tournament Selection |
| Crossover | Two-point crossover |
| Crossover probability | 0.9 |
| Mutation | Random flip |
| Mutation probability | 0.1 |

**Supplementary Table 3.** Sensitivity, specificity, and TPC for the four clinically inferred symptom combinations computed on the UK-discovery cohort.

| Symptom | Age group | 3-day analysis |  |  |  |  |  | 7-day analysis |  |  |  |  |  |
| --- | --- | --- | --- | --- | --- | --- | --- | --- | --- | --- | --- | --- | --- |
|  |  | Sensitivity (%) |  | Specificity (%) |  | TPC |  | Sensitivity (%) |  | Specificity (%) |  | TPC |  |
|  |  | UK | US | UK | US | UK | US | UK | US | UK | US | UK | US |
| Respiratory symptoms<sup>1</sup> | All | 44.7 | 48.1 | 81.9 | 76.6 | 43 | 21 | 57.7 | 64.6 | 79.1 | 72.3 | 38 | 19 |
|  | [18-54] | 45.2 | 45.2 | 81.6 | 76.8 | 38 | 20 | 58.2 | 64.3 | 78.4 | 71.6 | 33 | 18 |
|  | [55+] | 42.9 | 51.4 | 82.7 | 76.5 | 64 | 22 | 55.8 | 64.9 | 80.5 | 73.0 | 54 | 20 |
| WHO-defined pneumonia<sup>2</sup> | All | 58.8 | 74.7 | 71.9 | 59.5 | 51 | 23 | 70.3 | 84.8 | 68.5 | 54.5 | 46 | 23 |
|  | [18-54] | 59.2 | 69.0 | 71.1 | 61.3 | 45 | 22 | 70.5 | 83.3 | 67.5 | 55.5 | 41 | 21 |
|  | [55+] | 57.5 | 81.1 | 73.3 | 57.8 | 73 | 24 | 69.6 | 86.5 | 70.7 | 53.6 | 65 | 25 |
| COVID-19-specific symptoms<sup>3</sup> | All | 67.4 | 79.7 | 69.8 | 57.6 | 48 | 23 | 81.0 | 92.4 | 66.4 | 52.6 | 43 | 22 |
|  | [18-54] | 68.1 | 73.8 | 69.0 | 59.5 | 42 | 22 | 82.0 | 92.9 | 65.3 | 53.8 | 38 | 20 |
|  | [55+] | 65.1 | 86.5 | 71.5 | 55.7 | 69 | 24 | 77.7 | 91.9 | 68.8 | 51.4 | 62 | 25 |
| Extended Symptoms<sup>4</sup> | All | 90.7 | 96.2 | 26.0 | 21.1 | 86 | 35 | 95.5 | 98.7 | 23.0 | 17.9 | 82 | 35 |
|  | [18-54] | 91.2 | 95.2 | 25.0 | 22.2 | 75 | 32 | 95.5 | 100.0 | 21.7 | 18.6 | 72 | 32 |
|  | [55+] | 88.9 | 97.3 | 28.1 | 20.1 | 128 | 38 | 95.4 | 97.3 | 25.7 | 17.3 | 119 | 39 |

<sup>1</sup>Cough, dyspnoea; <sup>2</sup>Cough, dyspnoea, fever; <sup>3</sup>Fever, cough, dyspnoea, and anosmia/ageusia; <sup>4</sup>Fever, cough, dyspnoea, anosmia/ageusia, fatigue, and headache TPC = Tests per case

**Supplementary Table 4.** Pareto of optimal combination of symptoms for three-day analysis computed on the UK-discovery training data, ordered by decreasing sensitivity.

| Symptom combinations | Sensitivity | Specificity | TPC |
| --- | --- | --- | --- |
| fatigue, anosmia/ageusia, cough, diarrhoea, headache, sore throat | 93.8 | 11.8 | 88 |
| fatigue, fever, anosmia/ageusia, cough, headache, sore throat | 92.5 | 14.9 | 85 |
| fatigue, anosmia/ageusia, cough, dyspnoea, headache, sore throat | 92.2 | 15.3 | 85 |
| fatigue, anosmia/ageusia, cough, hoarse voice, headache, sore throat | 92.0 | 15.5 | 85 |
| fever, anosmia/ageusia, cough, diarrhoea, headache, sore throat | 91.7 | 17.8 | 82 |
| anosmia/ageusia, cough, diarrhoea, hoarse voice, headache, sore throat | 91.2 | 18.9 | 81 |
| fatigue, fever, anosmia/ageusia, cough, diarrhoea, headache | 90.6 | 22.4 | 78 |
| fatigue, anosmia/ageusia, cough, dyspnoea, diarrhoea, headache | 90.5 | 22.7 | 77 |
| fatigue, anosmia/ageusia, cough, chest pain, sore throat, unusual muscle pains | 89.9 | 24.0 | 76 |
| fatigue, anosmia/ageusia, cough, dyspnoea, sore throat, unusual muscle pains | 89.8 | 24.7 | 75 |
| fatigue, fever, anosmia/ageusia, cough, dyspnoea, headache | 89.4 | 26.1 | 74 |
| fatigue, fever, anosmia/ageusia, cough, delirium, headache | 88.6 | 26.9 | 73 |
| fatigue, fever, anosmia/ageusia, cough, headache | 88.4 | 27.3 | 73 |
| fatigue, anosmia/ageusia, cough, dyspnoea, headache | 87.7 | 27.7 | 72 |
| fever, anosmia/ageusia, cough, dyspnoea, diarrhoea, headache | 87.3 | 31.3 | 69 |
| anosmia/ageusia, cough, dyspnoea, diarrhoea, hoarse voice, headache | 87.0 | 31.7 | 68 |
| fever, anosmia/ageusia, cough, diarrhoea, delirium, headache | 86.7 | 31.9 | 68 |
| anosmia/ageusia, cough, diarrhoea, delirium, hoarse voice, headache | 86.1 | 32.2 | 68 |
| anosmia/ageusia, cough, dyspnoea, hoarse voice, headache, unusual muscle pains | 86.0 | 35.2 | 65 |
| fever, anosmia/ageusia, cough, dyspnoea, hoarse voice, headache | 85.8 | 35.8 | 64 |
| fever, anosmia/ageusia, cough, delirium, hoarse voice, headache | 85.1 | 36.1 | 64 |
| fever, anosmia/ageusia, cough, dyspnoea, delirium, headache | 84.9 | 36.4 | 64 |
| anosmia/ageusia, cough, dyspnoea, delirium, hoarse voice, headache | 84.6 | 37.2 | 63 |
| anosmia/ageusia, cough, hoarse voice, headache, unusual muscle pains | 84.2 | 37.4 | 63 |
| fever, anosmia/ageusia, cough, dyspnoea, headache | 84.1 | 38.3 | 62 |
| anosmia/ageusia, cough, dyspnoea, hoarse voice, headache | 83.7 | 39.2 | 61 |
| fatigue, fever, anosmia/ageusia, cough, dyspnoea, unusual muscle pains | 83.5 | 40.0 | 60 |
| fatigue, anosmia/ageusia, cough, dyspnoea, hoarse voice, unusual muscle pains | 83.2 | 40.3 | 60 |
| fatigue, anosmia/ageusia, cough, dyspnoea, chest pain, unusual muscle pains | 82.5 | 40.7 | 59 |
| fatigue, fever, anosmia/ageusia, cough, dyspnoea, delirium | 82.1 | 41.6 | 58 |
| fatigue, fever, anosmia/ageusia, cough, dyspnoea | 82.0 | 42.3 | 58 |
| fatigue, anosmia/ageusia, cough, dyspnoea, unusual muscle pains | 81.5 | 42.8 | 57 |
| fatigue, fever, anosmia/ageusia, cough, delirium | 80.6 | 43.4 | 57 |
| fatigue, fever, anosmia/ageusia, cough | 80.2 | 44.1 | 56 |
| fatigue, anosmia/ageusia, cough, unusual muscle pains | 79.4 | 44.7 | 55 |
| fatigue, fever, anosmia/ageusia, delirium, unusual muscle pains | 78.5 | 45.2 | 55 |
| fatigue, fever, anosmia/ageusia, unusual muscle pains | 78.2 | 45.9 | 54 |
| fatigue, fever, anosmia/ageusia, dyspnoea | 77.6 | 46.0 | 54 |
| fatigue, anosmia/ageusia, dyspnoea, unusual muscle pains | 76.9 | 46.7 | 53 |
| fatigue, fever, anosmia/ageusia, delirium | 76.1 | 47.7 | 52 |
| fatigue, fever, anosmia/ageusia | 75.6 | 48.5 | 52 |
| fever, anosmia/ageusia, cough, diarrhoea, chest pain, unusual muscle pains | 75.4 | 51.4 | 49 |
| fever, anosmia/ageusia, cough, dyspnoea, diarrhoea, unusual muscle pains | 75.0 | 52.4 | 48 |
| fever, anosmia/ageusia, cough, diarrhoea, delirium, unusual muscle pains | 74.2 | 53.1 | 47 |
| anosmia/ageusia, cough, dyspnoea, diarrhoea, hoarse voice, unusual muscle pains | 74.0 | 53.7 | 46 |
| fever, anosmia/ageusia, cough, dyspnoea, abdominal pain, unusual muscle pains | 73.8 | 53.8 | 46 |
| fever, anosmia/ageusia, cough, dyspnoea, diarrhoea, hoarse voice | 73.7 | 54.2 | 46 |
| fever, anosmia/ageusia, cough, dyspnoea, hoarse voice, unusual muscle pains | 73.5 | 58.5 | 42 |
| fever, anosmia/ageusia, cough, dyspnoea, chest pain, unusual muscle pains | 72.8 | 59.1 | 41 |
| fever, anosmia/ageusia, cough, dyspnoea, skipped meals, hoarse voice | 72.6 | 60.1 | 40 |
| anosmia/ageusia, cough, dyspnoea, chest pain, hoarse voice, unusual muscle pains | 72.1 | 61.3 | 39 |
| fever, anosmia/ageusia, cough, dyspnoea, unusual muscle pains | 71.1 | 62.6 | 37 |
| anosmia/ageusia, cough, dyspnoea, hoarse voice, unusual muscle pains | 70.2 | 64.8 | 35 |
| fever, anosmia/ageusia, cough, dyspnoea, hoarse voice | 69.2 | 65.4 | 35 |
| anosmia/ageusia, cough, dyspnoea, chest pain, unusual muscle pains | 68.1 | 65.6 | 34 |
| fever, anosmia/ageusia, cough, dyspnoea, chest pain | 67.9 | 65.9 | 34 |
| fever, anosmia/ageusia, cough, dyspnoea, delirium | 67.8 | 65.9 | 34 |
| fever, anosmia/ageusia, cough, unusual muscle pains | 67.6 | 66.9 | 33 |
| fever, anosmia/ageusia, cough, skipped meals | 66.4 | 69.0 | 31 |
| fever, anosmia/ageusia, cough, dyspnoea | 65.9 | 70.1 | 30 |
| fever, anosmia/ageusia, cough, delirium | 64.3 | 70.5 | 30 |
| fever, anosmia/ageusia, cough | 61.5 | 75.3 | 25 |
| fever, anosmia/ageusia, dyspnoea | 55.8 | 76.4 | 24 |
| fever, anosmia/ageusia, skipped meals | 55.3 | 76.6 | 23 |
| anosmia/ageusia, cough, hoarse voice | 53.9 | 79.3 | 21 |
| anosmia/ageusia, cough, dyspnoea | 53.2 | 79.5 | 21 |
| anosmia/ageusia, cough, delirium | 49.0 | 79.7 | 20 |
| fever, anosmia/ageusia | 47.5 | 83.7 | 16 |
| anosmia/ageusia, cough | 45.6 | 85.9 | 14 |
| anosmia/ageusia, hoarse voice | 35.5 | 87.1 | 13 |
| anosmia/ageusia, delirium | 24.6 | 89.2 | 11 |

TPC = Tests per case

**Supplementary Table 5.** Pareto of optimal combination of symptoms for seven-day analysis computed on the UK-discovery training data, ordered by decreasing sensitivity.

| Symptom combinations | Sensitivity | Specificity | TPC |
| --- | --- | --- | --- |
| fatigue, anosmia/ageusia, cough, diarrhoea, headache, sore throat | 97.5 | 93.8 | 90 |
| fatigue, fever, anosmia/ageusia, cough, headache, sore throat, | 96.7 | 91.6 | 87 |
| fatigue, fever, anosmia/ageusia, cough, diarrhoea, sore throat, | 96.0 | 87.3 | 82 |
| fatigue, fever, anosmia/ageusia, cough, sore throat, unusual muscle pains | 95.5 | 84.0 | 79 |
| fatigue, anosmia/ageusia, cough, dyspnoea, sore throat, unusual muscle pains | 95.0 | 83.7 | 78 |
| fatigue, anosmia/ageusia, cough, hoarse voice, sore throat, unusual muscle pains | 94.8 | 83.5 | 78 |
| fatigue, anosmia/ageusia, cough, delirium, sore throat, unusual muscle pains | 94.7 | 83.4 | 77 |
| fatigue, anosmia/ageusia, cough, sore throat, unusual muscle pains | 94.5 | 83.0 | 77 |
| fatigue, fever, anosmia/ageusia, cough, headache | 93.8 | 82.5 | 76 |
| anosmia/ageusia, cough, dyspnoea, diarrhoea, sore throat, unusual muscle pains | 92.8 | 77.8 | 71 |
| fatigue, fever, anosmia/ageusia, cough, dyspnoea, diarrhoea | 92.3 | 75.3 | 68 |
| fatigue, fever, anosmia/ageusia, cough, abdominal pain, unusual muscle pains | 92.2 | 74.3 | 67 |
| fatigue, fever, anosmia/ageusia, cough, dyspnoea, unusual muscle pains | 91.7 | 71.1 | 64 |
| fatigue, fever, anosmia/ageusia, cough, unusual muscle pains | 91.0 | 69.8 | 62 |
| fatigue, fever, anosmia/ageusia, cough, dyspnoea | 90.5 | 69.5 | 62 |
| fatigue, anosmia/ageusia, cough, dyspnoea, unusual muscle pains | 90.2 | 69.3 | 61 |
| fatigue, fever, anosmia/ageusia, persistent cough | 89.7 | 68.2 | 60 |
| fatigue, anosmia/ageusia, cough, unusual muscle pains | 89.2 | 68.1 | 59 |
| fatigue, fever, anosmia/ageusia, unusual muscle pains | 88.4 | 67.2 | 58 |
| fatigue, fever, anosmia/ageusia, skipped meals | 88.2 | 67.1 | 58 |
| fatigue, anosmia/ageusia, persistent cough | 87.0 | 66.5 | 57 |
| fatigue, fever, anosmia/ageusia | 86.9 | 65.4 | 55 |
| fever, anosmia/ageusia, cough, dyspnoea, diarrhoea, unusual muscle pains | 86.0 | 61.5 | 52 |
| anosmia/ageusia, cough, dyspnoea, diarrhoea, hoarse voice, unusual muscle pains | 85.9 | 60.1 | 50 |
| anosmia/ageusia, cough, dyspnoea, diarrhoea, skipped meals, unusual muscle pains | 85.4 | 59.8 | 50 |
| anosmia/ageusia, cough, dyspnoea, abdominal pain, hoarse voice, unusual muscle pains | 85.2 | 59.0 | 49 |
| fever, anosmia/ageusia, cough, dyspnoea, hoarse voice, unusual muscle pains | 85.0 | 54.8 | 45 |
| anosmia/ageusia, cough, dyspnoea, skipped meals, hoarse voice, unusual muscle pains | 84.5 | 54.1 | 45 |
| anosmia/ageusia, cough, dyspnoea, chest pain, hoarse voice, unusual muscle pains | 84.4 | 51.8 | 43 |
| anosmia/ageusia, cough, dyspnoea, hoarse voice, unusual muscle pains | 82.7 | 48.6 | 39 |
| anosmia/ageusia, cough, dyspnoea, chest pain, unusual muscle pains | 81.2 | 48.3 | 38 |
| fever, anosmia/ageusia, cough, unusual muscle pains | 79.7 | 47.5 | 37 |
| anosmia/ageusia, cough, dyspnoea, chest pain, hoarse voice | 79.5 | 44.1 | 34 |
| fever, anosmia/ageusia, cough, dyspnoea | 78.4 | 44.0 | 33 |
| anosmia/ageusia, cough, dyspnoea, hoarse voice | 76.5 | 40.0 | 30 |
| anosmia/ageusia, cough, unusual muscle pains | 75.4 | 39.4 | 29 |
| anosmia/ageusia, cough, dyspnoea, chest pain | 75.2 | 39.4 | 29 |
| fever, anosmia/ageusia, persistent cough | 73.9 | 39.0 | 28 |
| anosmia/ageusia, dyspnoea, unusual muscle pains | 72.4 | 38.6 | 27 |
| fever, anosmia/ageusia, dyspnoea | 72.2 | 38.1 | 27 |
| anosmia/ageusia, cough, dyspnoea | 71.9 | 34.1 | 24 |
| anosmia/ageusia, dyspnoea, chest pain | 66.6 | 34.0 | 22 |
| anosmia/ageusia, unusual muscle pains | 65.4 | 30.4 | 19 |
| fever, anosmia/ageusia | 64.6 | 29.9 | 18 |
| anosmia/ageusia, persistent cough | 64.1 | 26.8 | 16 |
| anosmia/ageusia, dyspnoea | 60.1 | 26.6 | 15 |
| anosmia/ageusia, delirium | 50.4 | 25.8 | 12 |

TPC = Tests per case

## Supplementary Figures

**Supplementary Figure 1.**
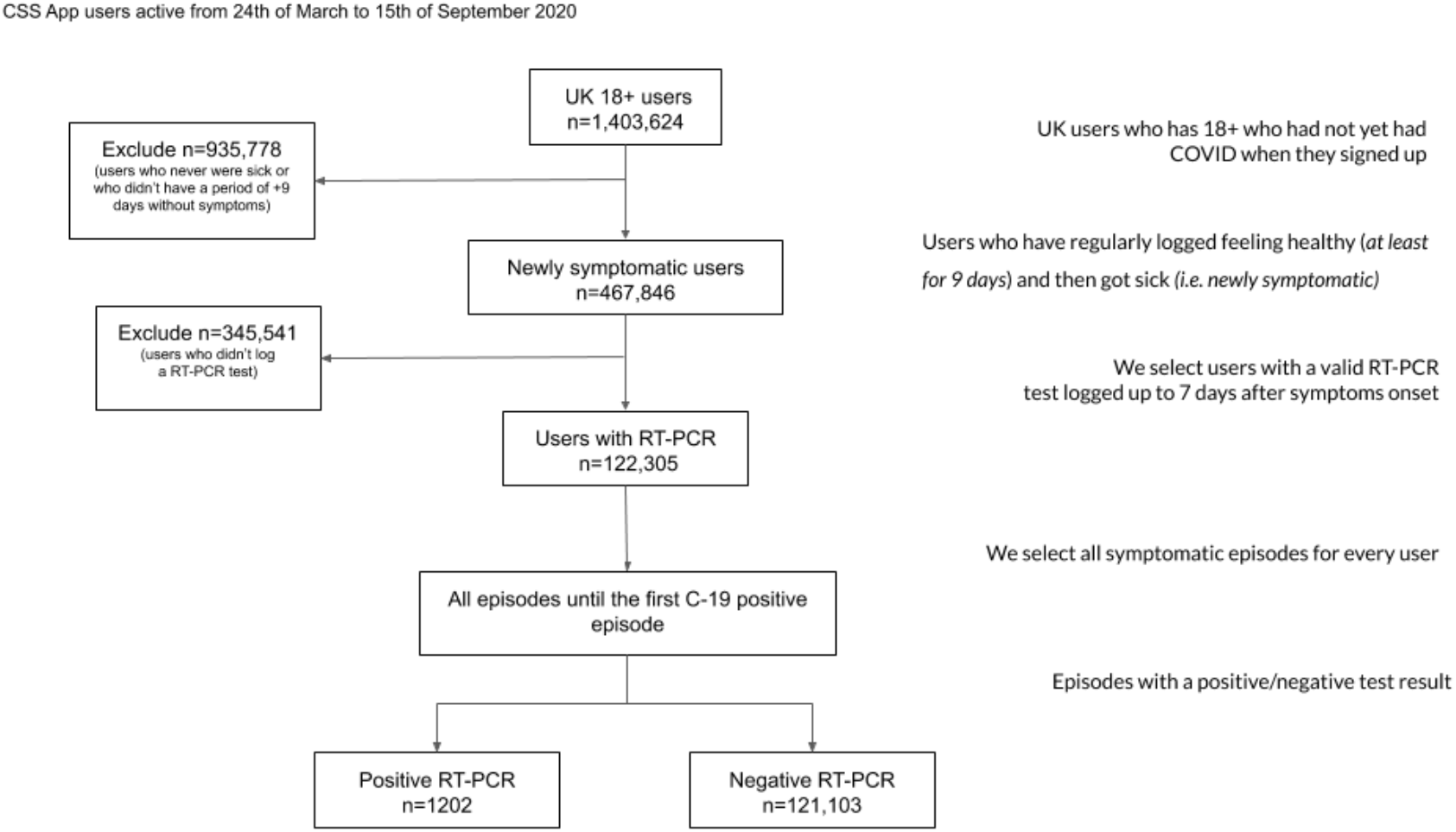
Flow diagram of user selection for the UK-discovery cohort.

**Supplementary Figure 2.**
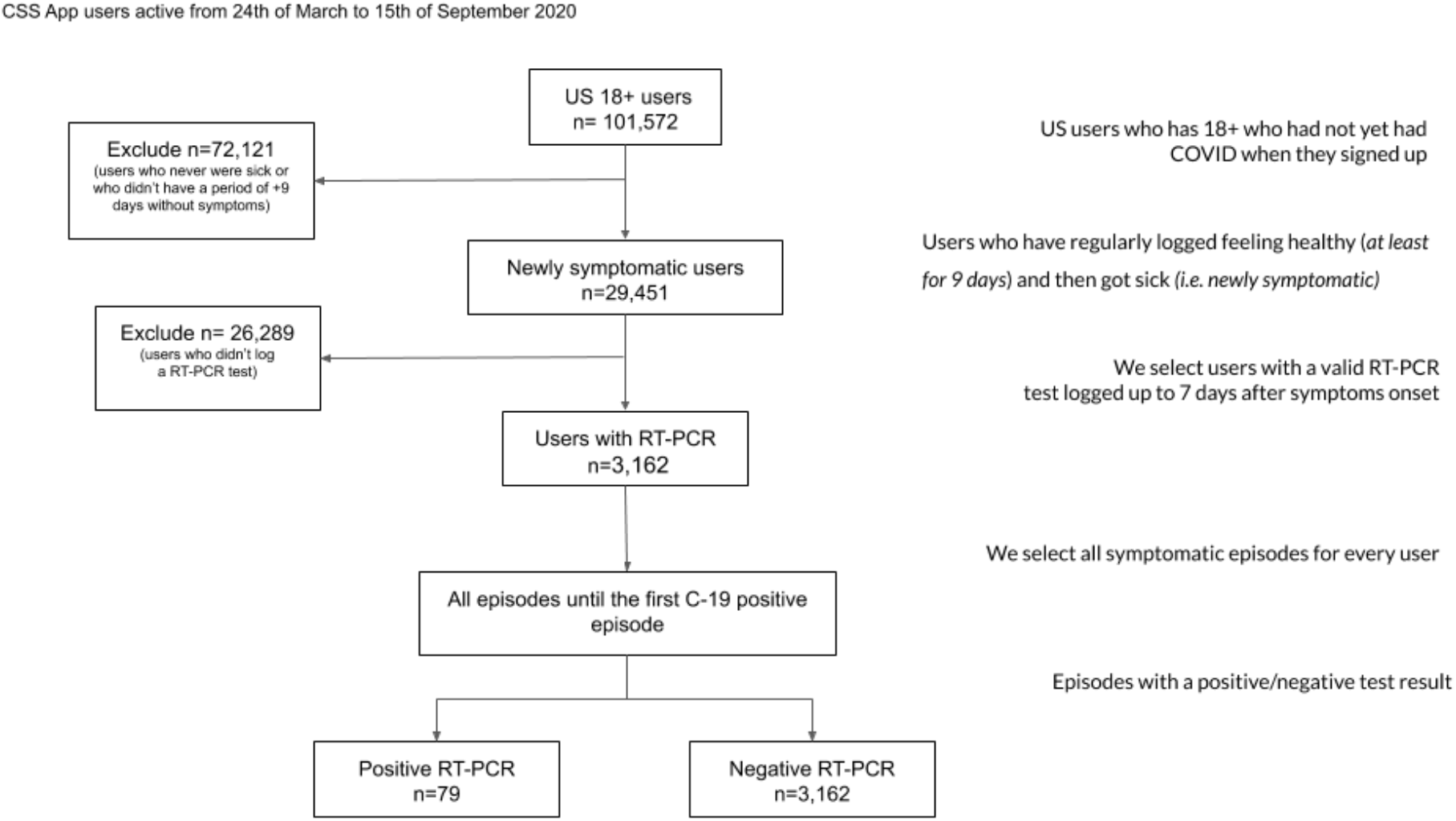
Flow diagram of user selection for the US-replication cohort.

